# An Accurate Model for SARS-CoV-2 Pooled RT-PCR Test Errors

**DOI:** 10.1101/2020.12.02.20242651

**Authors:** Yair Daon, Amit Huppert, Uri Obolski

## Abstract

PCR testing is an important tool to mitigate outbreaks of infectious diseases. One way of increasing testing throughput is by simultaneously testing multiple samples for the presence of a pathogen, a technique known as *pooling*. During the current COVID-19 pandemic, rapidly testing individuals for the presence of SARS-CoV-2 is conducted in large amounts. Since testing is often a bottleneck in mitigating the spread of SARS-CoV-2, pooling is increasing in popularity. Most analyses of the error rates of pooling schemes assume that including more than a single infected sample in a pooled test does not increase the probability of a positive outcome. We challenge this assumption with experimental data and suggest a novel probabilistic model for the outcomes of pooled tests. As an application, we analyze the false-negative rates of one common pooling scheme known as Dorfman pooling. We show that the false-negative rates of Dorfman pooling increase when the prevalence of infection decreases. However, low infection prevalence is exactly the condition under which Dorfman pooling achieves highest throughput. We therefore implore the cautious use of pooling and development of pooling schemes that consider correctly accounting for tests’ error rates.

## Introduction

Reverse transcription polymerase chain reaction (RT-PCR) testing is a key component in breaking transmission chains and mitigating the COVID-19 pandemic. As such, the need for large-scale testing has resulted in the development of techniques to increase the the throughput of RT-PCR tests [2, 8, 13, 14, 16]. These techniques, often termed *pooling*, or group testing, have originated in the seminal work of Dorfman and his pooling technique [2, 7]. Under Dorfman pooling, one selects *N* individuals and performs a single RT-PCR test on their combined (*pooled*) samples. If the pooled test yields a positive result, then each individual is retested separately; otherwise, everyone is declared negative. The throughput efficiency of Dorfman pooling has been demonstrated empirically [2] and its error rates thoroguhly investigated [1, 11, 17].

Studies focused on pooling for SARS-CoV-2 are in a consensus that sample dilution effects [10,21] are not a concern, even for pools as large as 64 individuals [2, 9, 14, 24]. Consequently, studies assumed that the probability of a true-positive (the test’s *sensitivity*) does not depend on the number of infected samples in the pool, but rather on the existence of at least one such sample. Thus, the probability of a positive result in a pooled test has been (assumed) identical for a pool with one sample from an infected individual and, e.g., five such samples. This assumption is common in the group testing literature [1, 11], as well as in more specific, COVID-19 focused studies [3, 17]. In this study we challenge this commonly made assumption and show how using a more accurate probabilistic model affects estimation of false-negative rates for Dorfman pooling.

## Methods

Formally, we consider a pool containing *N* individuals {1, …, *N*}. We denote the true infection state *θ* ∈ {0, 1}^*N*^, so individual *i* is infected iff *θ*_*i*_ = 1. The RT-PCR test’s sensitivity (true-positive rate) is denoted *S*_e_, and the test’s specificity (true-negative rate) is denoted *S*_p_. Pooled test result (data) is denoted **d** ∈ {0, 1}, where **d** = 0 iff the test returned a negative result.

### The common assumption

Previous studies of pooling schemes assumed that the false-negative probability does not depend on the number of infected samples, but merely on the existence of at least one such sample in a pool [1, 11]. Current studies of pooling in the context of SARS-CoV-2 also employ a similar assumption [3, 17]. Explicitly, these studies assume:

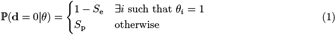

Below, we refer to (1) as the *common assumption*.

### Refuting the common assumption

We refute the common assumption with experimental data collected from [6], and summarized in Table 1. There, the authors investigate Dorfman pooling and, regardless of the pooled test result, follow up and test each pool member separately. We focus on 128 pools for which at least one subsequent separate test was positive — of which 29 pooled tests were negative and 99 positive. In the data cited in [6], of the 29 negative pools, subsequent separate testing yielded a single positive result in 24. In contrast, of the 99 positive pools, 42 yielded a single positive test upon subsequent separate testing.

**Table 1:** Contingency table of data from [6]

|  | Negative pool | Positive pool |
| --- | --- | --- |
| # subsequent positives = 1 | 24 | 42 |
| # subsequent positives $\geq 1$ | 5 | 57 |
| Total | 29 | 99 |

The data in Table 1 allows us to test the following null hypothesis *H*_0_: The probability of a pooled false-negative is equal for pools with one subsequent positively tested member and pools with two or more such members. *H*_0_ is a direct consequence of the common assumption, and rejecting *H*_0_ implies the common assumption is not realistic, at least for SARS-CoV-2.

We apply Fisher’s exact test for the presence of more than one positive individual in correctly identified pools. Fisher’s test yields an increased odds ratio of 6.4, 95% CI (2.2,23.4), with a p-value ≈ 10^−4^. Thus, we reject *H*_0_, refuting the common assumption.

### Our model

Since the essence of the refuted common assumption is that amplification of all samples occurs only once, we assume a more realistic model: amplification of viral RNA succeeds or fails for each sample independently. Furthermore, according to [1, 3, 11, 17], a false-positive does not depend on the number of negative samples in a pool. For lack of data pointing otherwise, we incorporate this assumption into our model with a small modification. We do assume that a false amplification can occur only once per pool. However, we also assume false amplification is independent of any other correct amplification. Specifically, it is possible that every correct amplification fails *and* an erroneous one occurs simultaneously. This assumption is somewhat specific for the current application of screening for SARS-CoV-2 via RT-PCR. For example, cross-reactivity with other coronaviruses would have violated this assumption. However, cross-reactivity was ruled out in [19]. These assumptions lead to the following model, which is illustrated in Figure 1 and summarized in (2).

**Figure 1:**
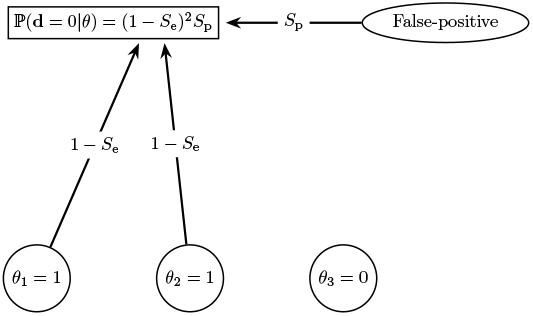
Illustration of the revised probabilistic model. A pool contains individuals {1, 2, 3} with state *θ* = (1, 1, 0) (i.e., individuals 1 and 2 are infected). A negative pooled test (**d** = 0) occurs when three detection paths fail: A false-negative occurs for individuals 1 and 2, each with probability (1 −*S*_e_). Additionally, no false-positive detection occurs, with probability *S*_p_. Individual 3 is not infected and does not contribute to the probability of the pooled test result.

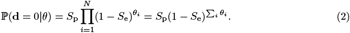

### Application: scheme false-negative rate

We calculate the false-negative rate for a single *infected* individual, henceforth referred to as “Donald”, under our model and under the common assumption. We distinguish three types of false-negative events when performing pooling. A *single test*’s false-negative is the event of a negative result upon testing Donald separately, i.e., in an RT-PCR test without pooling. A *pooled* false-negative occurs when a pooled test containing Donald’s sample (and other samples) yields a negative result, i.e., the pooling fails to detect at least one positive result. Lastly, a *scheme* false-negative occurs when an entire pooling scheme fails to identify Donald as infected. Our first task is to calculate Dorfman’s scheme false-negative rate. Equivalently, we ask: what is the probability of not identifying Donald as infected under Dorfman pooling?

Denote the prevalence of infection in the (tested) population *q*. We denote Donald as individual 1, so that *θ*_1_ = 1. Then:

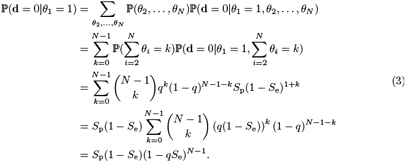

If the pooled test yields a positive result, Donald is tested separately. Taking a conservative stand, it is assumed that such a simple procedure poses no risk of introducing contaminant RNA. Therefore, the separate test yields a positive result with probability *S*_e_.

We calculate the probability that Donald is mistakenly identified as not infected, henceforth referred to as the scheme’s false-negative rate and denoted *S*_fn_. In order to correctly identify an infected individual as infected, both pooled and separate tests have to yield a positive result. Thus, the scheme’s false-negative rate is:

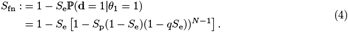

### Comparison metric

The single test false-negative rate 1 − *S*_e_ and scheme false-negative rate *S*_fn_ are compared via:

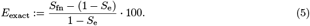

*E*_exact_ is the percentage increase in the pooling scheme false-negative rate, relative to the single test false-negative rate.

According to the common assumption, the scheme false-negative rate is %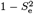. A straight forward calculation shows that this implies the percentage increase in scheme false-negative rate is *E*_common_ := 100 *· S*_e_.

## Results

### Scheme false-negative

We plot *E*_exact_ for varying prevalence *q* and sensitivity *S*_e_ values, and make the comparison with *E*_common_. As recommended by [2], we apply different pool sizes *N*, for different prevalence values. We observe that for a false-positive rate *S*_p_ = 0.95 [2] and a range of reasonable sensitivity and prevalence values [19, 20, 22, 23], an increase of at least 60% in *E*_exact_ can be expected (Figure 2).

**Figure 2:**
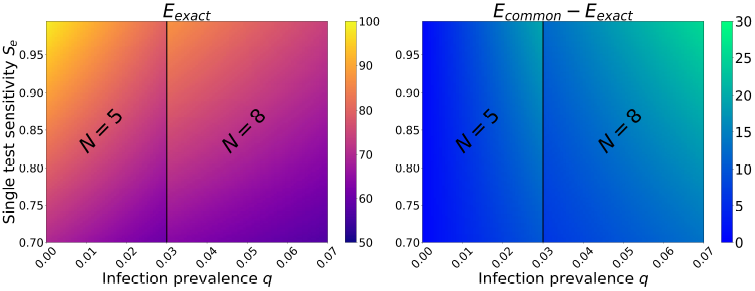
Relative increase in Dorfman pooling false-negative rates *E*_exact_. Left: Colors represent *E*_exact_, the relative percentage increase in the scheme false-negative rates relative to the single test false-negative rates (5). Right: colors represent the difference between *E*_common_ and *E*_exact_. The disease prevalence *q*, is varied on the x-axis, while the test sensitivity is varied on the y-axis. Pool size *N*, was chosen according to *q* as in [2]. The left panel shows that *E*_exact_ is largest for low prevalence values *q*. The difference between *E*_common_ and *E*_exact_ can be as large as 30%, as seen in the right panel.

Interestingly, an increase in infection prevalence monotonically decreases the scheme false-negative rate, as can also be easily seen from (4). For the chosen parameter ranges, the increase in the single test false-negative rates increases the relative error *E*_exact_. These effects can be seen in Figure 2 (left panel), upon conditioning on pool size. Extending the range for *S*_p_ yields no qualitative differences. We further compare *E*_exact_ to *E*_common_, showing the discrepancy changes as a function of both prevalence and the single test sensitivity (Figure 2, right panel).

## Discussion

Here, we developed a more realistic and novel probabilistic model for the outcomes of pooled PCR tests, parametrized for SARS-CoV-2. Contrary to the common assumption, we assume, based on data (Table 1), that multiple infected individuals increase the likelihood of a positive pooled test result. A direct consequence of our model is that false-negative rates depend on infection prevalence. Specifically, low values of infection prevalence increase the false-negative rates of Dorfman pooling. These results remain qualitatively similar under varying parameter values, in the observed ranges [12, 19, 20, 22] (Figure 2). Our results give rise to a conflict: low infection prevalence leads to high efficiency of Dorfman pooling [2], while also increasing false-negative rates.

As the COVID-19 pandemic progresses, the infection prevalence in various tested populations undergoes frequent changes. Hence, as our results suggest, pooling schemes employed for mass testing should be used with caution in populations where infection rates are low. Such mass-tested populations often include air travel passengers [5] or presymptomatic and asymptomatic individuals [18], and can be crucial for controlling outbreaks [15].

Thus, improving pooling schemes is imperative. An especially important consideration in designing such schemes is explicitly taking the intrinsic PCR error rates into account. We [4], as well as others [16], have developed such models, although they have not yet been implemented in real-world settings.

To conclude, pooling is an important technique that can increase testing throughput in a cost-effective manner. Nevertheless, care must be given to pooling schemes’ false-negative rates, especially under low infection prevalence settings.

## Data Availability

Data was extracted from the paper Sample pooling for SARS-CoV-2 RT-PCR screening by De Salazar et al. cited in our paper.

## Acknowledgements

Yair Daon was supported by a post-doctoral fellowship from the Tel Aviv University Center for Combating Pandemics and the Raymond and Beverly Sackler dean’s post-doctoral fellowship.

